# Epidural stimulation restores muscle synergies by modulating neural drives in participants with motor/sensory complete spinal cord injuries

**DOI:** 10.1101/2022.06.15.22276156

**Authors:** Rajat Emanuel Singh, Aliya Ahmadi, Ann Parr, Uzma Samadani, Andrei V. Krassioukov, Theoden I Netoff, David P. Darrow

## Abstract

Multiple studies have corroborated restored volitional motor control after motor-complete spinal cord injury (SCI) through the use of spinal cord stimulation (SCS/eSCS) but rigorous quantitative descriptions have been lacking. Using a structured surface electromyogram based (sEMG) task with and without SCS during the Epidural Stimulation After Neurological Damage (ESTAND) study in participants with chronic, motor and sensory complete SCI, we investigated muscle activity complexity and muscle synergies to better characterize neuro-muscular control.

In addition, competition exists between the task and neural origin hypotheses underlying muscle synergies, and this analysis in humans with motor and sensory complete chronic injury provided an opportunity to test these hypotheses. Muscle activity complexity was computed with Higuchi Fractal Dimensional analysis (HFD), and muscle synergies were estimated using non-negative matrix factorization (NNMF) in six participants with AIS A chronic SCI. We found that the complexity of muscle activity is immediately reduced with SCS in the SCI participants. We also found that over the follow-up sessions, the muscle synergy structure of the SCI participants became more defined, and the number of synergies decreased over time, indicating improved coordination between the muscle groups. Lastly, we found that the muscle synergies were restored with SCS, supporting the neural hypothesis of muscle synergies. We conclude that SCS restores muscle movements and muscle synergies that are distinct from healthy, able-bodied controls.

## Introduction

More than 800,000 people suffer a traumatic SCI every year worldwide (*Kumar et al., 2018*), with about half resulting in motor and sensory complete paralysis (*Cells, 2017*). Patients who have not regained motor control after one year rarely go on to do so and are considered to have chronic SCI (*Kirshblum et al., 2004*). Exercise or activity-based therapies (locomotor and non-locomotor training) with functional electrical stimulation, which rely on residual ascending pathways (*Mushahwar et al., 2007*), (*Lam et al., 2007*), (*Craven et al., 2017*), (*Marquez-Chin and Popovic, 2020*), remain the most effective treatments in such cases, especially for incomplete SCI (*Jones et al., 2014*). However, individuals with complete SCI (no motor or sensory function below the level of the injury) lack the neural control or feedback necessary to benefit from these therapies to train and restore voluntary movement.

The ESTAND clinical study has shown that epidural stimulation in patients with chronic and complete SCI (AIS A score) can restore voluntary motor function (*Pino et al., 2020*), (*Darrow et al., 2019*). To measure the recovery of motor control, we employed a standardized sEMG based brain motor control assessment (BMCA) with and without stimulation. Changes in volitionally controlled muscle activity with and without stimulation are observed immediately after turning on the stimulator. Changes in motor control due to tonic neuromodulation and measured during the BMCA task afforded us an opportunity to measure neuroplasticity in humans. Despite ongoing optimization, we previously reported that the total sEMG activity of the legs seemed to plateau and even decrease after the first six months despite a subjective improvement in motor control (*Pino et al., 2020*), (*Zhao et al., 2021*). As a result, we endeavored to objectively characterize changes in neuromuscular control over time with SCS therapy associated with spinal cord plasticity to better elucidate this discrepancy.

We propose to use muscle synergy analysis (*Singh et al., 2018*) and complexity analysis (*Santuz et al., 2018*), (*Santuz et al., 2020*) to quantify changes in sEMG patterns during participants’ recoveries. Muscle synergies are considered to have a modular organization in the CNS, which when activated by neural drives forms a movement. Thus, synergies and the associated neural drives can explain neurophysiological characteristics of a movement (*Singh et al., 2018*). Furthermore, we map muscle activity to the spinal cord (rostro-caudal plane) to estimate how activation within the spinal cord changes with stimulation over time (*Ivanenko et al., 2006*).

There are two competing hypotheses on the origin of muscle synergies, one suggesting a neural basis and the other a task-dependent basis. The task-based synergy hypothesis states that the task determines the synergies; thus, changes in the synergies reflect changes in the dynamics of the task, limb biomechanics, and/or musculoskeletal structure (*Kutch and Valero-Cuevas, 2012*), (*Cheung and Seki, 2021*). The neural synergy hypothesis states that changes in these synergies (number and structure of muscle synergies) reflect neuromodulatory changes directly mediated by the CNS (*Singh et al., 2018*), (*Bizzi and Cheung, 2013*). Several studies have tested and validated these hypotheses using participants with and without movement disorders (*Abd et al., 2021b*). However, it has been very difficult to disambiguate the two hypothesized origins of these synergies in humans because it is challenging to do the same task in participants with and without neural control. SCS in participants with motor and sensory complete SCI provides a unique opportunity to complete the same task under both conditions by modulating the neural control of the local spinal circuitry, which allows us to directly address the origin of muscle synergies. We hypothesized that muscle synergies have neural origin and believe this is the first study to provide direct evidence for their neural basis in humans. In addition, this framework allows us to examine the effect of long-term epidural stimulation on muscle synergies during the recovery of participants with complete motor and sensory SCI, which we hypothesized would demonstrate improvements but with distinct divergence from able-bodied control participants.

## Material & Methods

### Participant recruitment/description

This study has been approved by the Hennepin Healthcare Research Institute Institutional Review Board with an Investigational Device Exemption from the United States Food and Drug Administration. We analyzed six participants with motor and sensory complete SCI (AIS A), (*Kirshblum et al., 2011*) who completed at least 7 follow-up sessions. The demographic and medical information of each participant is listed in Table 1. The injuries for all SCI participants were between spinal levels T4 and T8. All SCI participants were implanted with an epidural stimulator consisting of a three-column, 16-contact paddle lead through a T12-L1 laminectomy, and an internal pulse generator (IPG) with a primary cell (Tripole and Proclaim Elite, Abbott, Plano, TX, United States) was placed subcutaneously in the lower lumbar area under general anesthesia, as shown in Figure 1. Follow-up visits were performed monthly for up to one year (13 follow-ups). SCI participants from the ESTAND study that had completed at least 7 of the follow-up sessions were included in this analysis. A detailed description of the study can be found in previous publications (*Darrow et al., 2019*), (*Pino et al., 2020*), (*Pino et al., 2022*). In addition, nine healthy participants were also recruited to undergo the BMCA as controls for this study. At each follow-up session after surgery, SCI participants underwent a BMCA with and without stimulation (*Pino et al., 2020*).

**Table 1:** Demographic data of participants

| Pat. ID | Age (Decade) | AIS Score | Injury Level | Time Since Injury (Years) | Sessions |
| --- | --- | --- | --- | --- | --- |
| SCI001 | 30s | A | T4 | 8 | 13 |
| SCI002 | 40s | A | T8 | 18 | 13 |
| SCI003 | 50s | A | T5 | 6 | 13 |
| SCI004 | 60s | A | T5 | 4 | 13 |
| SCI005 | 30s | A | T4 | 9 | 7 |
| SCI006 | 20s | A | T6 | 2 | 10 |

**Figure 1:**
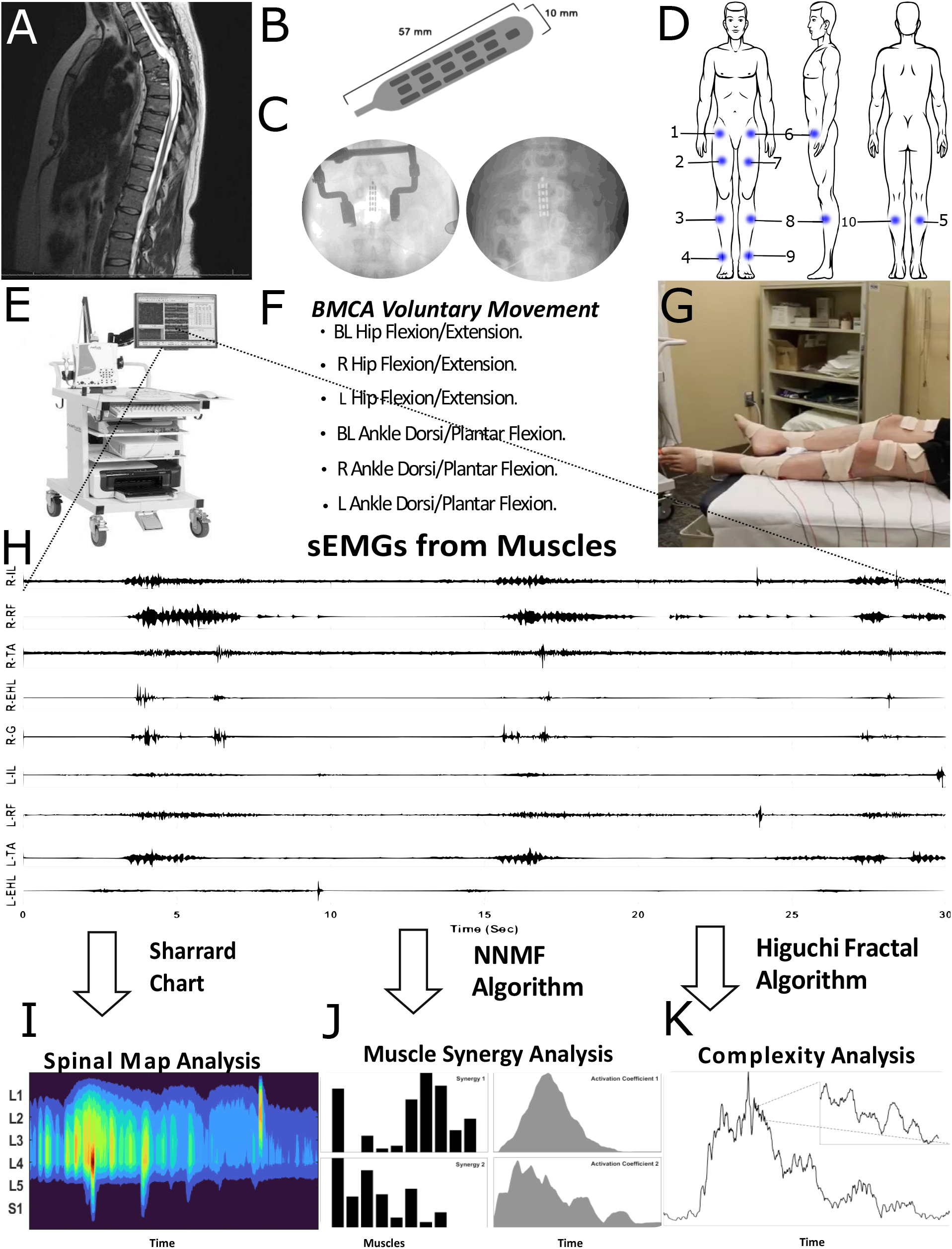
Experimental design and protocol. A) CT scan showing a SCI. B) Abbott TripoleTM 16-contact lead. C) X-rays of leads implanted in SCI participants; Left: paddle implanted during T12 laminectomy surgery; Right: paddle implanted after surgery overlying the T12-L1 epidural space. D) Left: front view; Middle: side view; and Right: back view showing the placement of EMG electrodes in blue. Electrodes were placed on the 1. Right Iliopsoas (R-IL), 2. Right Rectus Femoris (R-RF), 3. Right Tibialis Anterior (R-TA), 4. Right Extensor Hallucis Longus (R-EHL), 5. Right Gastrocnemius (R-G), 6. Left Iliopsoas (L-IL), 7. Left Rectus Femoris (L-RF), 8. Left Tibialis Anterior (L-TA), 9. Left Extensor Hallucis Longus (L-EHL), and 10. Left Gastrocnemius (L-G). E) Integrated Computer-Nicolet EDX EMG system powered by viking software used to acquire sEMGs during the BMCA. F) BMCA tasks performed by the SCI participants and control participants after electrode placement while in the supine position (G). H) Sample sEMGs, which were analyzed to understand changes in the neuro-muscular control using spinal map analysis, muscle synergy analysis, and fractal analysis (I, J, K, respectively).

### BMCA protocol

The BMCA task is an electrophysiologic assessment of voluntary motor control that involves relaxation, reinforcement maneuvers (deep breath, neck flexion, Jendrassik maneuver, and bilateral shoulder shrug), and voluntary leg movements (bilateral hip flexion/extension, isolated hip flexion/extension of left and right side, bilateral ankle dorsiflexion/plantarflexion followed by isolated dorsiflexion/plantarflexion of left and right foot) (*Sherwood et al., 1996*), (*Pino et al., 2020*).

In each trial, a two-toned auditory cue sounded twice to signal the control participants and SCI participants to begin and end the movement. Three trials of each voluntary movement were performed by following the two-tone auditory cue played three times. For example, after hearing the first tone, the participants would flex their hips, and after the second tone, the participants would extend their hip, and end the movement. Three trials were performed for each voluntary movement.

For the control participants, the BMCA protocol was conducted once. For the SCI participants, the complete BMCA protocol was conducted twice at each follow-up visit, once with stimulation and again without stimulation. If the SCI participants were not able to perform the movement, they were asked to follow the protocol and perform as they were able. During each follow-up session, three trials of data were acquired with and without stimulation for all six voluntary movements for a total of 36 trials = 2 conditions × 3 trials × 6 voluntary movements. For the control participants, we acquired 18 trials = 1 condition × 3 trials × 6 voluntary movements.

### EMG recording, processing, and segmentation

sEMG recordings of the 10 lower limb muscles were acquired during the BMCA tasks at a sampling rate of 600 Hz using the Nicollet EDX EMG system from the right and left sides of the following muscles: iliopsoas (R-IL, L-IL), rectus femoris (R-RF, L-RF), tibialis anterior (R-TA, L-TA), gastrocnemius (R-G, L-G), and extensor hallucis longus (R-EHL, L-EHL).

MATLAB (2020b, Natik MA) was used to process and analyze the sEMG data. Each channel was filtered using a 6th-order bandpass Butterworth filter from 10 Hz to 300 Hz. For muscles in close proximity to the stimulation electrode and thus having strong stimulus artifacts, such as the R-IL and L-IL, a 5th-order median filter was used to remove the stimulation artifact. Instantaneous power, *sEMG*_RMS_, was estimated by calculating the root mean square (RMS) envelope over 100-millisecond non-overlapping windows.

The beginning and end of each movement, as indicated by auditory tone, were labeled with timestamps in the EMG acquisition system. These timestamps were used to segment the voluntary movement *sEMG*_RMS_ for each trial. Because each trial was of different length, the segmented *sEMG*_RMS_ signals were time-normalized by interpolation to 7000 time points, resulting in an *sEMG*_RMS_ matrix size of 3 trials × 10 channels × 7000 time points.

sEMG processing was performed for each voluntary movement (VM = 1 to 6) under the complete BMCA protocol (condition × trials × [channel × time points] × VM = 1 × 3 × [10 × 7000] × 6) for control participants and (2×3×[10 × 7000]×6) for SCI participants (with and without stimulation conditions).

## Complexity Analysis

We used HFD analysis on the *sEMG*_RMS_ to quantify the complexity in the *sEMG*_RMS_ amplitude over time (Higuchi 1988; Cukic et al. 2018). *sEMG*_RMS_ was analyzed over time as a sequence of N samples, *E*(1), *E*(2), … *E*(*N*), and k new time series sequences were created.

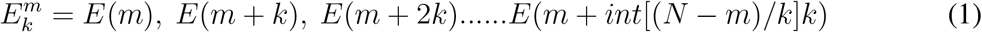

In this equation, the initial time (m) = 1,2,3 … .k, the time interval (k) = 2, 3,..… *k*_max_, and int is the integer part of the real number. The length *L*_*m*_(*k*) of every time-series sequence was constructed using equation (2).

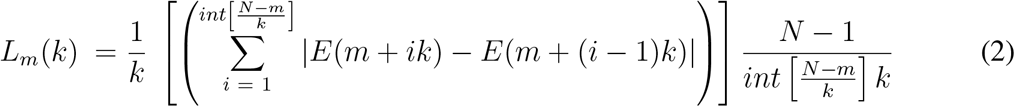

*L*_*m*_(*k*) is averaged across all m, resulting in an average curve length of *L*_*k*_, which is computed from equation (3). Moreover, the HFD was determined from the slope of the best fit of *ln*(*L*_*k*_) vs. 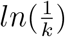 using equation (4).

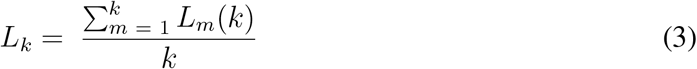

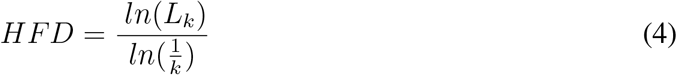

HFD analysis was performed on each muscle during the voluntary movements in the BMCA task. The HFD complexity was measured with and without stimulation for every voluntary movement and muscle over the follow-up sessions. We compared the HFD complexity of the control participants with that of the SCI participants with and without stimulation. In addition, a non parametric test was performed on the complexity values of the SCI participants’ muscles over the follow-up sessions with and without stimulation.

### Spinal motor output

We also studied the effect of epidural stimulation on spinal cord activity using maps of muscle activity. We mapped the *sEMG*_RMS_ onto the estimated rostro-caudal region of the motor neuron (MN) pool in the spinal cord from segments L1 to S1 (*Santuz et al., 2020*), (*Ivanenko et al., 2006*). A myotomal chart developed by Sharrard and shown in equation (5) was used to calculate the maps of putative alpha motor neuron activation (*Ivanenko et al., 2006*), (*Sharrard, 1964*). The myotomal chart provides the connection between each muscle and a specific spinal segment. The chart models muscle innervation from the spinal segments via the alpha motor neurons. The activity in the spinal cord was computed from a SCI participant’s voluntary movement with and without stimulation.

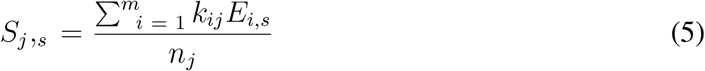

Here, *S*_*j,s*_ is the estimated spinal motor output from the jth segment with s samples, *E*_*i,s*_ is the *sEMG*_RMS_ signal from channel/muscle i, m is the number of sEMG signals, *k*_*ij*_ is the weighting coefficient of the ith muscle corresponding to the jth segment, and *n*_*j*_ is the number of *sEMG*_RMS_ values to the jth segment. The weighting coefficient *k*_*ij*_ values in the spinal maps are based on those from previous studies (*Sharrard, 1964*), (*Kendall et al., 1993*), (*Ivanenko et al., 2006*).

### Muscle synergy extraction

Muscle synergies are considered to be organized in the CNS as low-dimensional muscle coactivation patterns used to form movement (*Bizzi et al., 1991*), (*Bizzi and Cheung, 2013*). In short, muscle synergies are the functional building blocks of movement extracted from sEMG linear envelopes. Muscle synergies and their activation coefficients are generally estimated using non-negative matrix factorization (NNMF) (*Singh et al., 2018*), and they can explain the neurophysiological characteristics of a movement. We used the NNMF algorithm to estimate muscle synergies from the *sEMG*_RMS_ (*Lee and Seung, 1999*). The *sEMG*_RMS_ for each segmented trial was amplitude-normalized between 0 and 1, scaling from the minimum to the maximum recorded value. The segmented trials within each voluntary movement were ensemble-averaged, forming an *sEMG*_RMS_ matrix for each volitional movement of size (Muscles × timepoints = 10 × 7000). The NNMF algorithm was applied to this matrix to estimate muscle synergies. A mathematical model for time-invariant muscle synergies is given by equation (6).

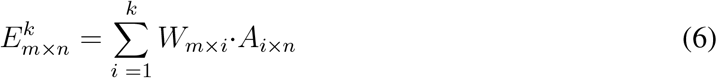

Here, *E*_*k*_ is the *sEMG*_RMS_ signal reconstructed with k extracted synergies, m is the number of muscles/channels, n is the number of samples/time points, W is the synergy (spatial structure), and A is the activation coefficient (temporal structure).

NNMF uses the multiplicative update rule method developed by (*Lee and Seung, 1999*). Hence, for synergy extraction, it was run 100 times to avoid local optima. Moreover, determining the number of synergies, k, is not trivial (*Singh et al., 2018*), (*Abd et al., 2021a*); therefore, prior to synergy extraction, we first determined the number of factors/synergies that explain 85% or more of the total variance, as calculated in equation (7).

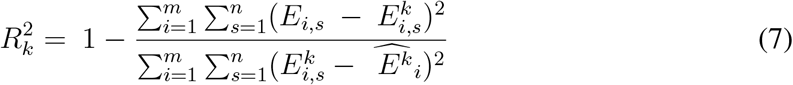

Here, *E*_*i,s*_ is the actual *sEMG*_RMS_ signal, 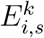 is the reconstructed *sEMG*_RMS_, 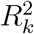 is the total variance explained by the first k components, and *Ê* is the mean of the reconstructed *sEMG*_RMS_.

The number of factors defined in the NNMF was selected to range from 1 to 10, where 10 is the maximum number of synergies that can be extracted, as determined by the number of muscle groups recorded.

The *sEMG*_RMS_ signals were reconstructed for each NNMF factor (1-N), and the *R*^2^ value for each defined synergy/factor was computed and plotted using equation (7). Based on previous studies, we defined k as the value for which 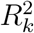 meets a minimum threshold of 85% (*Cheung et al., 2020*).

The muscle synergies were extracted for SCI participants’ voluntary movements during stimulation and without stimulation over the follow-up sessions. The control participants’ muscle synergies were also extracted during their single visit. The control participants’ muscle synergies were used to understand how muscle synergies change over follow-up sessions with stimulation, as we have previously observed that long-term eSCS improves volitional movement (*Pino et al., 2020*).

### Comparison of muscle synergies and their activation coefficients

To understand the impact of stimulation on muscle synergies, we compared the *R*^2^ curves and structures of muscle synergies (muscle loadings within the synergy vector) with and without stimulation during the BMCA. The *R*^2^ curves for control participants and SCI participants were compared for each voluntary movement performed during the BMCA. Moreover, the effects of stimulation on the *R*^2^ curves and muscle synergy structures over the follow-up sessions were also studied.

After extraction of the muscle synergies, the synergies were compared, sorted, and reordered based on similarity. We used the cosine similarity equation (8) to compare the muscle synergy structures. An R value close to 1 is considered highly similar, whereas an R value close to 0 suggests independence. Correlations between synergies across all participants were measured. The participant whose synergies had the highest correlation to those of the other participants was used as the template. All other participants’ synergies were then ordered to best match the template synergies for further analysis.

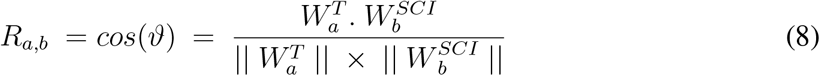

Here, 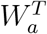 is the template synergy and 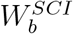 is the synergy for all other SCI participants compared to the template. a and b (1,2,…k,) are the number of synergies compared for SCI participants. The same procedure was implemented to organize the synergies of the control participants.

To compare the activation coefficients of respective muscle synergies, a zero-lag cross-correlation was used. A cross-correlation value close to 0 suggests a weak correlation, and a cross-correlation value close to 1 or -1 suggests a strong positive or negative correlation, respectively.

### Statistical analysis

To determine if the data were normally distributed, a Kolmogorov-Smirnov test was used. We used Mann Whitney U as a non-parametric test for non-normally distributed data. For statistical analysis, p *<* 0.05 was considered statistically significant.

Violin plots are used to display the differences between the mean, the median value of the control participants, and the SCI participants’ muscle activities with and without stimulation over the follow-up sessions.

## Results

### Epidural stimulation restores independent muscle activity

We first studied the effect of stimulation on independent muscle activity during BMCA tasks. Visual inspection of the sEMG waveforms indicated the restoration of muscle activity in the SCI participants during SCS. However, compared to the control participants, their sEMG waveforms were not as strong or concise in time, and localized to the specific muscle groups required to complete the task.

Figure 2 shows example sEMG profiles for a control participant and a SCI participant performing unilateral hip and ankle movements (right hip flexion/extension and right ankle plantar/dorsiflexion). During unilateral movement, the SCI participant showed increased sEMG amplitude on not only the ipsilateral side due to stimulation but also for the majority of muscle groups on the contralateral side. In the control participants, only the muscles directly involved in the task showed a higher sEMG amplitude. During the BMCA tasks, we found that stimulation evoked increased muscle activity with some asymmetry in the SCI participants. The asymmetric activation was likely a result of involuntary contraction of muscles on contralateral side.

**Figure 2:**
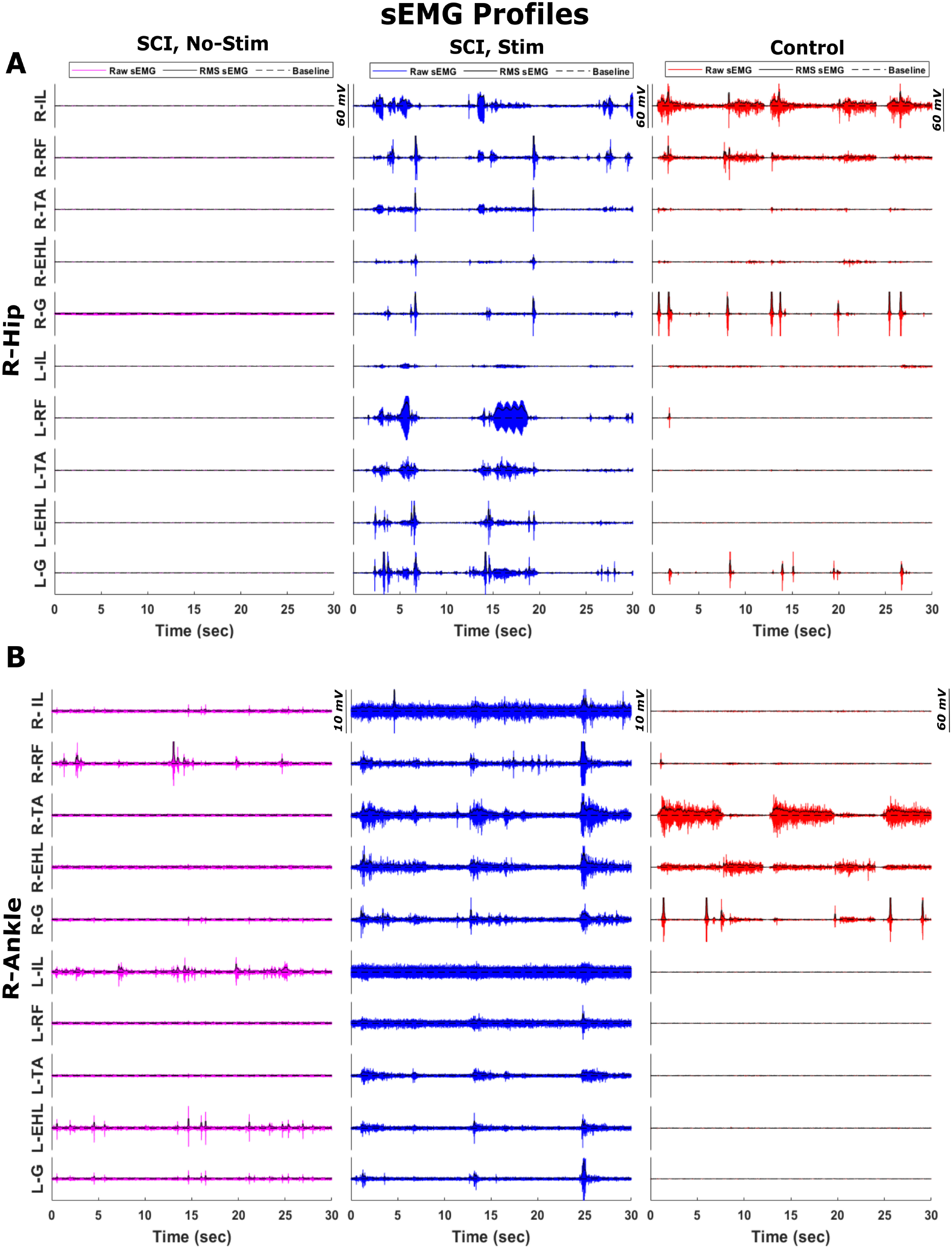
Raw sEMG profiles and their RMS envelopes obtained during BMCA tasks for SCI participants and controls. Examples of sEMG profiles obtained during BMCA tasks involving A) right hip and B) right ankle movement by SCI participants without stimulation (left column), SCI participants with stimulation (middle column), and control participants (right column). An RMS envelope calculated over 100 msec is also indicated. The zero offset is indicated by the dashed line.

### Muscle activation profile complexity

We also studied the effect of stimulation on the complexity of the *sEMG*_RMS_. The complexity of muscle movement is measured using the HFD, which detects the patterns and smoothness of muscle movement (*Müller et al., 2017*). Figure 3 shows violin plots of the fractal dimensions for control participants and SCI participants, with and without stimulation over the follow-up sessions. In SCI participants with stimulation, the HFD of muscle activity during the BMCA tasks was reduced significantly (p *<* 0.05, Mann-Whitney U-test) compared to that of SCI participants without stimulation. With stimulation, the HFD median values for muscle activation were close to those of the control participants, as shown in Figure 3. The lower HFD values across SCI participants during stimulation suggest that the complexity of muscle activation decreases as a result of eSCS. Moreover, unlike control participants, the reduced complexity of muscle activation for SCI participants with stimulation was not dependent on the task. In the control participants, muscles involved directly with the task had lower fractal dimensions than those not directly involved, indicating that selective activation of these muscles reduces the complexity.

**Figure 3:**
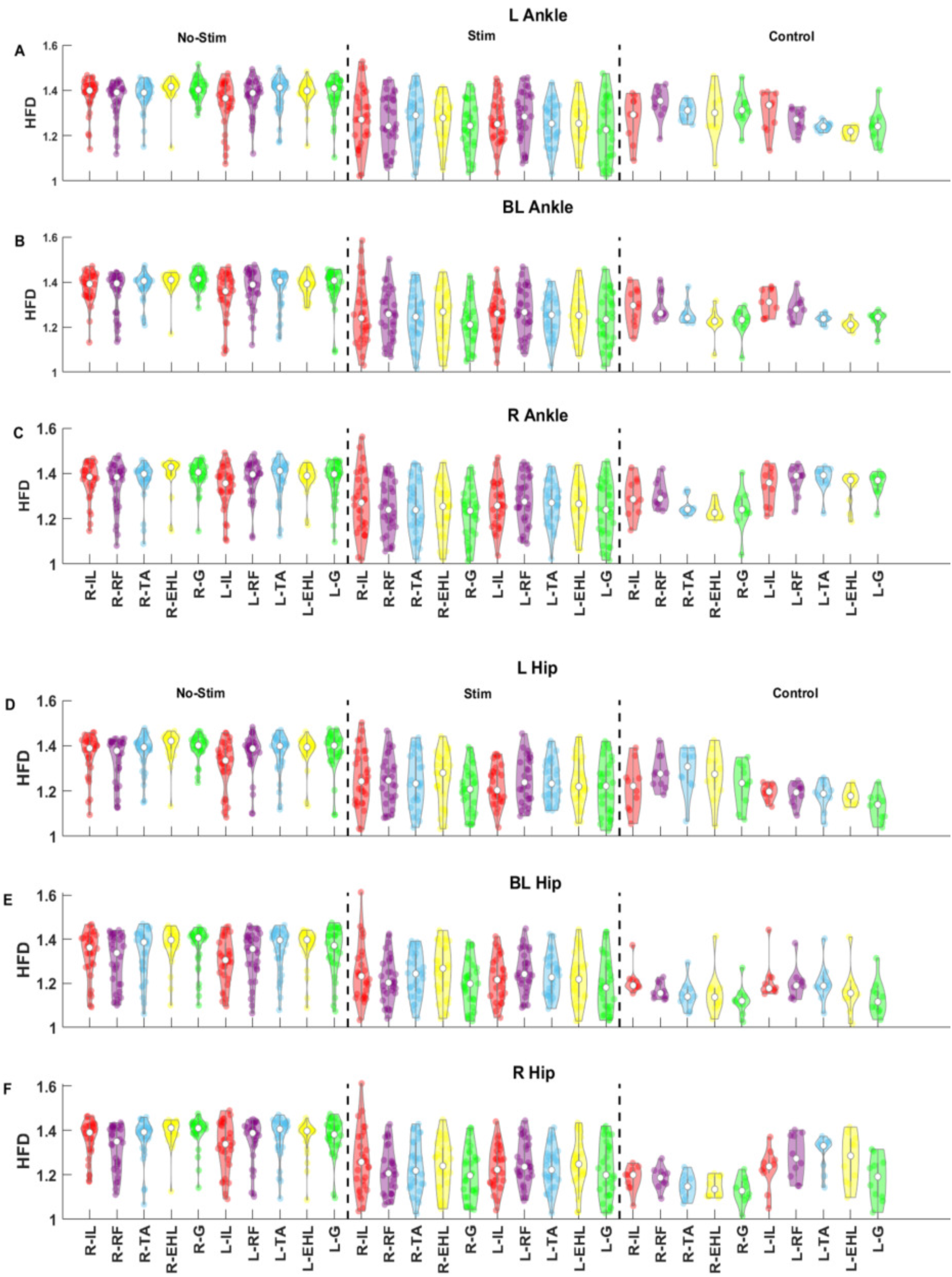
Higuchi Fractal Dimension (HFD) plots of sEMGs. HFD is used to measure the complexity of *sEMG*_RMS_ signals. The top panels (A, B, C) display ankle movement during the BMCA task, and the bottom panels (D, E, F) correspond to hip movement. The top rows in each panel (A, D) represent left ankle and hip movements, the second rows (B, E) represent BL ankle and hip movements, and the bottom rows (C, F) represent right ankle and hip movements. Within each row, the HFD is displayed for SCI participants without stimulation (left), SCI participants with stimulation (middle), and control participants (right). The control participants have much lower complexity than the SCI participants without stimulation. With stimulation of the SCI participants, the complexity decreases. In the control participants, the complexity is lower in the muscles involved in the task than in the muscles on the contralateral side.

### Estimated spinal motoneuron activity based on muscle activation

We then studied the effect of epidural stimulation on spinal cord activity by mapping the *sEMG*_RMS_ on the rostral-caudal plane of the spinal cord during BMCA tasks. This mapped spinal activity provided an estimate of alpha motor neuron activity. We first compared the mapped spinal activity (averaged over all trials) of each participant with and without stimulation and later compared the activity with those of the control participants. Figure 4 shows the averaged mapped spinal activities of the control and SCI participants with and without stimulation during bilateral movements. The estimated alpha motor neuron activity amplitude from each segment was significantly different between no stimulation and stimulation conditions in the SCI participants (p *<* 0.01, Mann-Whitney U-test). In addition, Figure 4 depicts localized estimated motor neuron activity in the temporal domain due to stimulation, suggesting that the mapped spinal activity is sensitive to epidural stimulation in the temporal domain.

**Figure 4:**
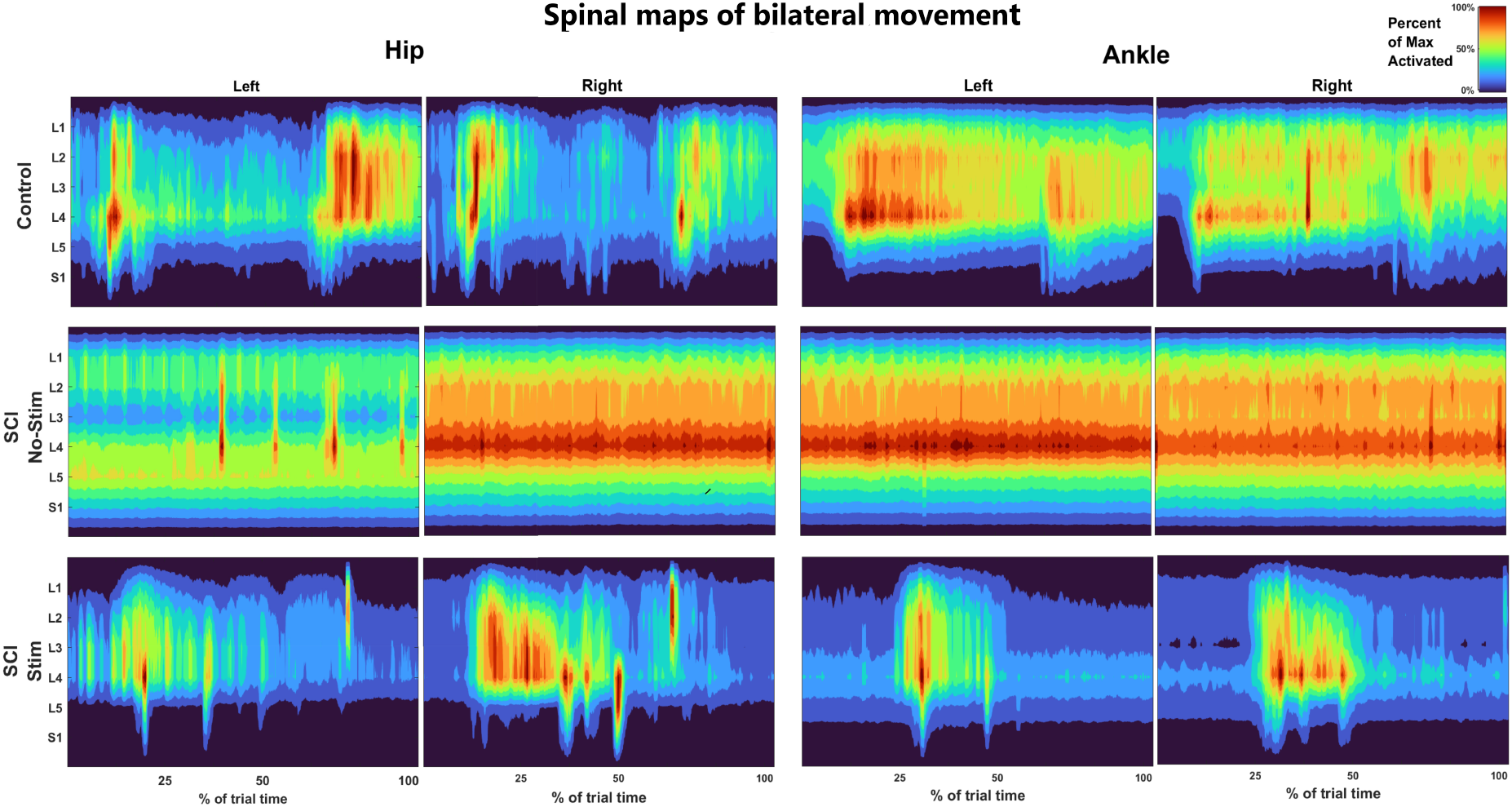
Spinal activity mapped during BL hip and BL ankle movements during BMCA in SCI participants and control participants. The x axis corresponds to movement as a percentage of task completion (temporal domain); the y axis depicts the spinal segment in the rostro-caudal plane from L1-S1 (spatial domain); and the heatmap represents the estimated alpha motor neuron activity. These estimated spinal activation patterns were obtained by mapping each muscle activation onto the relevant spinal segment based on the Sharrard chart (*Sharrard, 1964*). The left columns are the spinal activation patterns generated from muscle activity during BL hip movement. The right columns are the spinal activation patterns generated from muscle activity during BL ankle movement. The spinal activation patterns reveal that the estimated spinal activity is more localized in the temporal domain under stimulation conditions than in the control. The localization of spinal activity during BL hip movement suggests improper hip extension, as motor neurons are active mostly in the first half of contraction. The observed localization during BL ankle movement suggests reduced control as the tibialis anterior and gastrocnemius are co-contracted.

The difference in estimated alpha motor neuron activity amplitude for each segment between the control and SCI participants is also statistically significant (p *<* 0.001). From Table 2, most SCI participants showed localized motor neuron activity in a single phase (either at the beginning or the end of the trial) during BMCA tasks. On the other hand, the controls exhibited clearly separated activation events during the flexion and extension phases of the trial, as shown in Figure 4. In Figure 4, the controls do not show the clear localization of estimated motor neuron activity during ankle movement that was seen with the SCI participants. Hence, the estimated alpha motor neuron activity of SCI participants with stimulation is more localized in the temporal domain than the estimated alpha motor neuron activity of control participants.

### Muscle coordination improves with SCS therapy

We observed that the coordination capabilities of muscle groups during movement significantly improved with stimulation and over time, as measured by a decrease in the number of synergies. Figure 5 shows the *R*^2^ curves of SCI participants with and without stimulation and of the control subjects. The slopes of the *R*^2^ curves increase faster for hip movement than for ankle movement with stimulation over the follow-up sessions (1^*st*^, 5^*th*^, and last session), suggesting earlier restoration of the muscle synergies associated with hip movement than with ankle movement.

**Figure 5:**
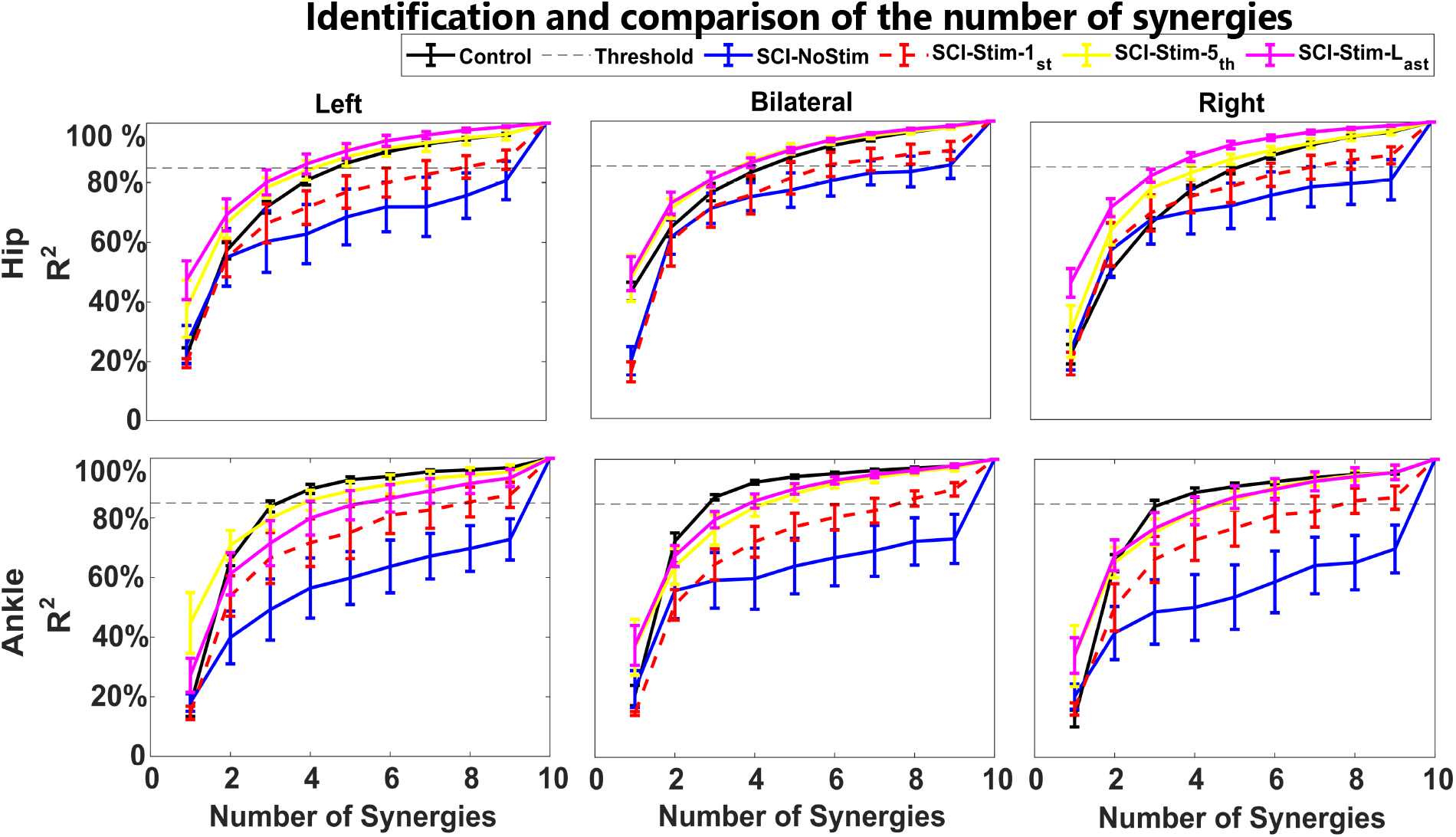
Number of muscle synergies determined from coefficient of determination (*R*^2^) curves across different therapeutic conditions. The number of synergies is determined from a threshold of 85% of the *R*^2^ value. The *R*^2^ value (y axis) is plotted against the number of factors/synergies (x -axis) for SCI participants under different conditions (no stimulation and stimulation in the 1st session, 5th session, and last session) and control participants. The left, middle, and right panels in the top row display left, bilateral, and right hip movements. The left, middle, and right panels in the bottom row display left, bilateral, and right ankle movements. In the absence of stimulation, all ten components were required to explain at least 85% variability in the data. During eSCS, a dose-dependent reduction in dimensionality was observed. In the last session, the number of synergies for the SCI participants was very close to that for the control participants. We found that a total of four muscle synergies were needed to explain 85% of the variance in the data–the same number of synergies needed for the control participants.

Figure 6 shows the muscle loadings for the extracted synergies up to the number of synergies for no stimulation, the first session with stimulation, and the final session with stimulation during bilateral hip and ankle movement tasks. As the number of synergies decreased, the structure of the muscle synergies also changed, as indicated by changes in the muscle loading values within a synergy during no stimulation, the first session with stimulation, and the final session with stimulation. Without stimulation, each muscle loading represented an individual muscle, indicating that the activation of each muscle was independent, and thus, no synergies were found. In the first session with stimulation, the muscle loadings increased, and few synergies were observed, where multiple muscles had significant loadings in the same synergy. In the final session, the muscle loadings were much stronger, and the synergies showed coordination between several muscles, with several muscles having high loadings within the same synergy and only 4 synergies being needed to explain 85% of the variance. These synergy loadings support our hypothesis that epidural stimulation restores muscle synergies by modulating both the structure and number of synergies.

**Figure 6:**
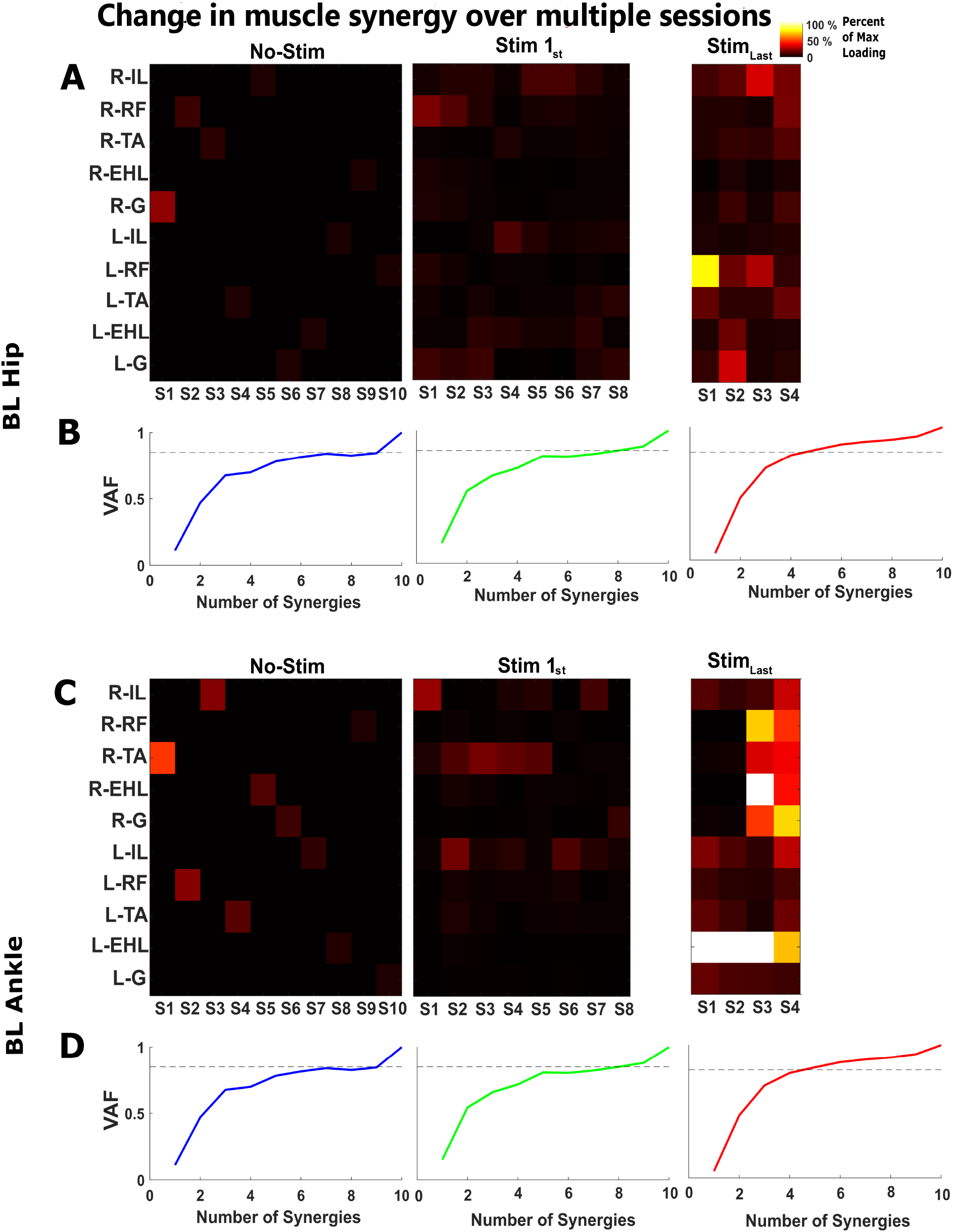
Spatial and temporal changes in muscle synergies. (A,C) Plots of muscle loading in which the x axis displays the synergies (S1, S2, S3—Sn) and the y axis displays the muscle groups. Each pixel in (A,C) is a muscle contribution/loading value within a synergy. (B,D) *R*^2^ curves; the dashed line is 85% of the total variance and is used as a threshold value for identifying the number of muscle synergies. We extracted ten, eight, and four muscle synergies based on the *R*^2^ threshold value across SCI participants without stimulation and with stimulation (1st and last session). Besides differences in the number of synergies, the muscle synergy structures (muscle loading values in a factor) of bilateral movements also changed between the first and last stimulation session. Relative to the first day of stimulation, the last day exhibited higher muscle loadings within a synergy and a smaller synergy space. Thus, both the structure and number of muscle synergies changed with eSCS.

### Muscle synergies shared across SCI participants

Based on the ordering scheme described in the methods, we calculated the R values across SCI participants for each BMCA task and ordered similar muscle synergies. The synergies extracted for SCI participants (last session) and control participants were compared, as the synergies in the last session exhibited dimensions/numbers consistent with those of the control participants. We found that the first four synergies were similar across SCI participants for each BMCA task, as shown by the high correlation (R values) depicted in Figure 7. This result indicates that the coordination of flexors and extensors was restored. However, temporal activation, which is likely determined by cortical control, is not directly affected by stimulation, and so the temporal patterns were not restored, as evidenced by weak correlations (R values near zero). In deafferented animal models, synergies are preserved but temporal patterns are weakened (*Cheung et al., 2005*) (*Cheung et al., 2012*), suggesting the central organization of synergies with a neural source (*d’Avella and Bizzi, 2005*), (*Singh et al., 2018*). Therefore, the restoration of similar synergies but not the temporal patterns across SCI participants with stimulation supports the hypothesis that these synergies originate from a neural source and not a task source.

**Figure 7:**
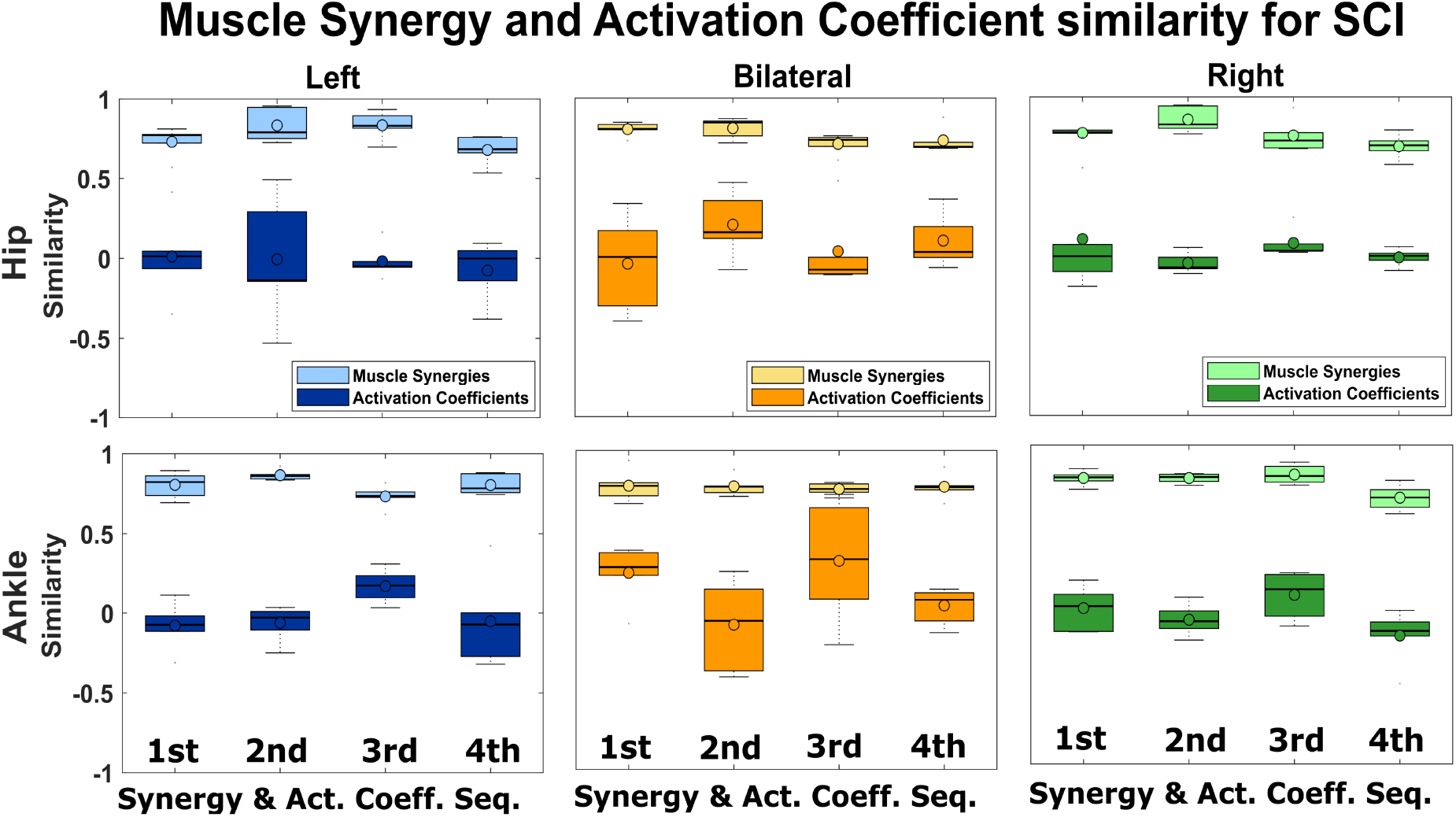
Comparison of synergies and activation coefficients across SCI participants. We observed four muscle synergies that were similar across SCI participants with epidural stimulation. The R values are plotted for the synergies and their activation coefficients across SCI participants. In the top row, the left, middle, and right panels display left, bilateral, and right hip movements, respectively. In the bottom row, the left, middle, and right panels show left, bilateral, and right ankle movements, respectively. The R values were computed using cosine similarity for the muscle synergies and zero-lag cross-correlation for the activation coefficients. Muscle synergies are more consistent than the respective activation coefficients across SCI participants.

### Muscle synergies of SCI participants vs control participants

The distinctively higher muscle loading (muscle loading *>* 0.5) values observed within a synergy among SCI participants and control participants (*Rimini et al., 2017*) display specific biomechanical functions. Therefore, we compared the muscle loadings of SCI participants with those of control participants. Figure 8 and Figure 9 show the muscle synergies of SCI participants and control participants.

**Figure 8:**
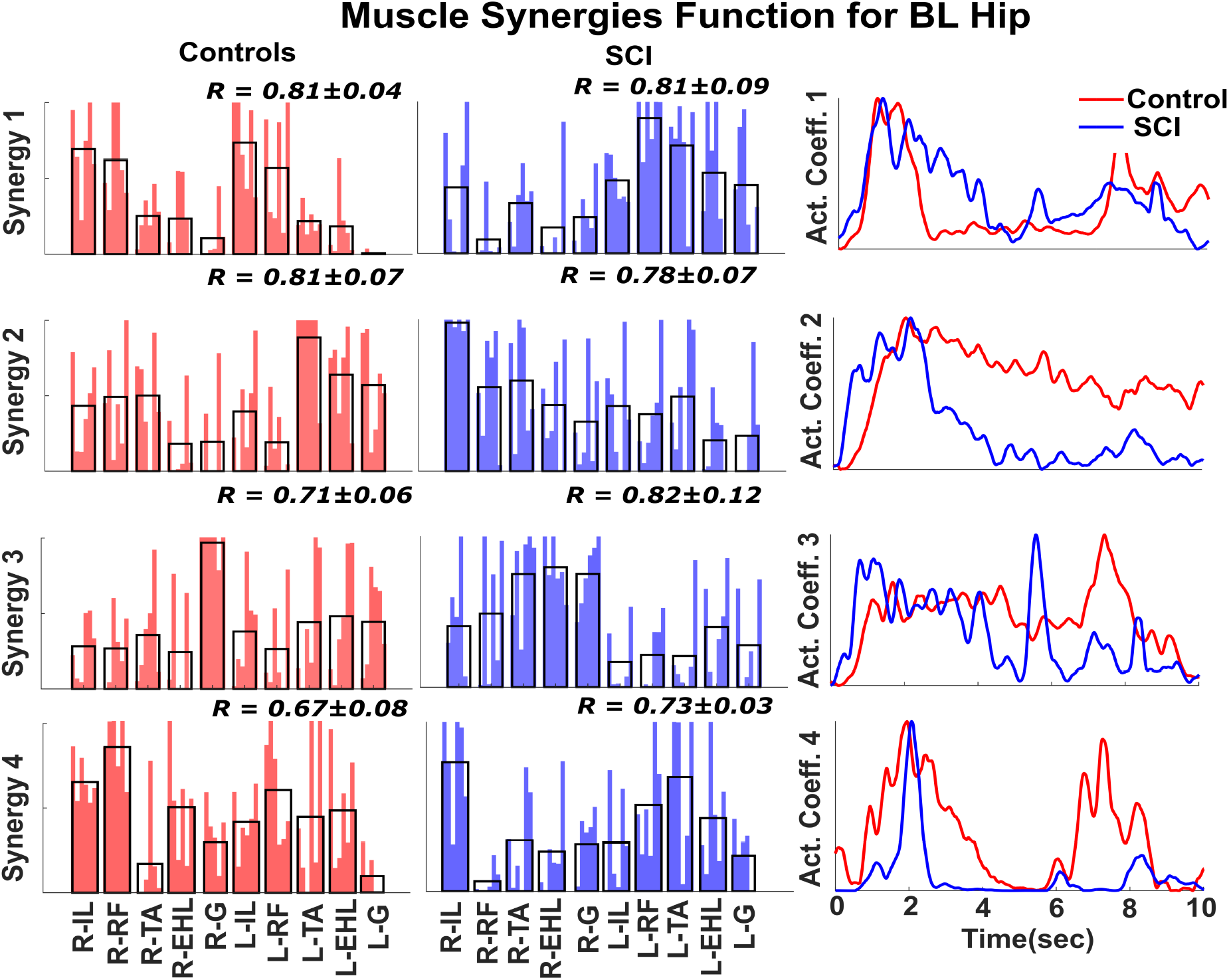
Muscle synergies for BL hip movement. Four similar synergies were identified across the control participants (left column) and SCI participants (middle column) during BL hip flexion. The weighting for each muscle synergy is shown for SCI participants and control participants. The average across weightings is shown as a black overlapping bar. In the right column, the activation coefficient is displayed as an average across SCI participants and control participants. Comparison of the BL hip movement synergies of SCI participants with those of control participants reveals asymmetry in the hip flexor muscle activity. The activity of the hip flexor muscles of the left and right sides is split into two separate synergies, synergy 1 and 2, while in control participants, synergy 1 includes activity for both sides.

**Figure 9:**
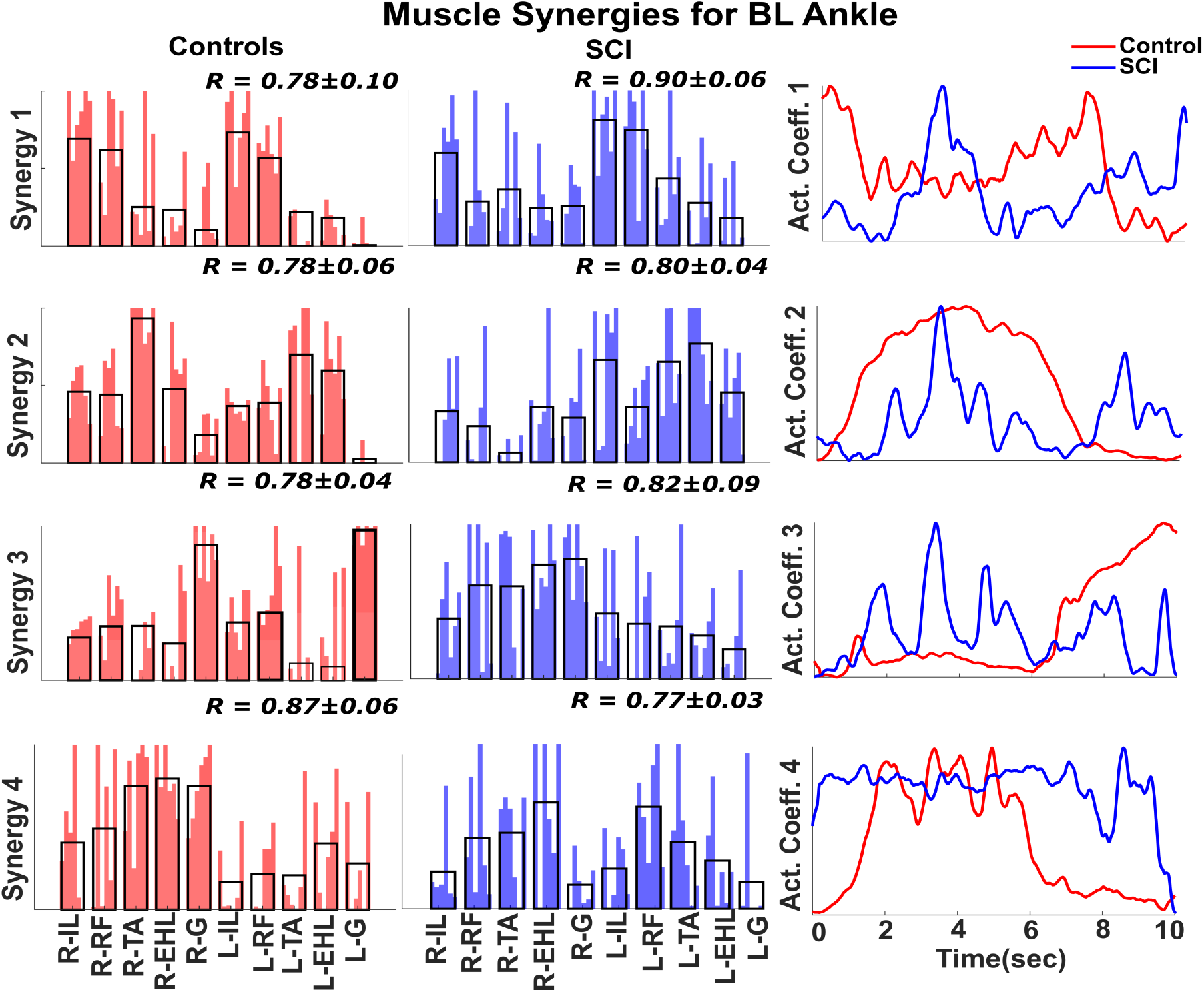
Muscle Synergies for BL ankle movement. Similar to the previous figure, synergies associated with BL ankle movement are plotted across control participants and SCI participants. The muscle synergies for BL ankle movement of the SCI participants also revealed asymmetric muscle contributions. The left- and right-side plantar flexor and dorsiflexor muscles are isolated in two synergies, 2 and 3, whereas for the control participants, a single synergy contains contributions from both sides’ dorsiflexor and plantar flexor muscles.

The first synergy observed during BL hip movement in control participants was different from that observed in SCI participants, as seen in Figure 8. In the control participants, the loadings in Synergy 1 were balanced between the left and right sides, indicating synergy between the two legs during bilateral movement. However, in the SCI participants, Synergy 1 represented left leg muscles and Synergy 2 represented right muscles, indicating a lack of synchronization between the legs during bilateral movement. Moreover, the muscle loadings in the remaining synergies of the SCI participants (Synergy 3 and Synergy 4) and control participants (Synergy 2 and Synergy 3) predominantly represented distal muscles, indicating that they may be associated with supporting knee flexion and foot inversion during BL hip flexion.

We found that the muscle synergies for BL ankle movement in SCI participants were also asymmetric. In control participants, the muscle loadings associated with left and right plantar flexion are seen in Synergy 2, and those of left and right dorsiflexion are seen in Synergy 3, as shown in Figure 9. In SCI participants, the synergies are asymmetric between the left and right sides, where Synergy 1 is more consistent with isometric hip flexion and Synergy 4 with foot inversion.

## Discussion

We studied the effect of eSCS on neuromuscular control in participants with motor and sensory complete, chronic SCI. We examined sEMGs acquired from the lower limbs of SCI participants while they performed a motor control task (BMCA) with and without stimulation over several follow-up visits and compared the results with sEMGs acquired from healthy participants. With stimulation, we observed a decreased complexity in muscle activation profiles over time in the follow-up visits. In addition, during the flexion or extension phase of BMCA tasks, SCI participants with stimulation exhibited more localized motor neuron activity than the control participants, as inferred from their muscle activation. We also studied the muscle synergies of the SCI participants to understand the modulation of neuromuscular control through stimulation in several follow-up sessions; over time, the muscle synergies showed dimensional and spatiotemporal changes. In particular, the number of muscle synergies decreased and the muscle loadings within the synergies increased between the first and last sessions. In comparison to the control participants, the muscle loadings of the SCI participants were asymmetric between the left and right sides. Overall, our results suggest that epidural stimulation modulates the local spinal circuitry to restore muscle synergies. However, these muscle synergies, while improved, remained significantly different from those of the control participants.

### Epidural stimulation and motor control

SCS, eSCS has been shown to restore some volitional control in patients with motor complete SCI. It is hypothesized that epidural stimulation activates local sensory afferents and motor efferents, which modulates the balance of excitation and inhibition in the spinal cord to restore a dynamic state that is responsive to the remaining supraspinal signals, thereby restoring volitional and autonomic control (*Darrow et al., 2019*), (*Eisdorfer et al., 2020*). Several studies have reported that epidural stimulation in conjunction with simple activity-based therapies restores voluntary control of movement (*Eisdorfer et al., 2020*). The therapies range from static postures, such as standing with full weight bearing, to simple dynamic movements, such as assisted stepping (*Harkema et al., 2011*). Moreover, epidural stimulation after incomplete SCI has restored volitional movement control for complex dynamic tasks such as treadmill walking (*Courtine et al., 2009*), (*Beck et al., 2020*). Recent studies have suggested that the epidural stimulation parameters can be optimized further for in-the-loop (*Zhao et al., 2021*), closed-loop, and/or phasic stimulation (*Formento et al., 2018*) to achieve more efficient voluntary control of movement.

In previous studies, we have shown that SCS results in increased sEMG amplitudes and restored movement (*Darrow et al., 2019*), (*Pino et al., 2020*). In this study, we further show that stimulation changes the complexity of muscle activation and activity patterns in the spinal cord. Spinal cord activity was estimated from muscle activity through a mapping procedure to estimate the alpha motor neuron activity in the dorsal roots (*Ivanenko et al., 2006*). The complexity of the movements was measured through HFD analysis of muscle activity. The mapped spinal activity and complexity analysis performed in this study identified differences in the control of movement between SCI participants and control participants. The mapped spinal activity of SCI participants indicated co-contraction of the distal muscles during bilateral movement and involuntary contraction of contralateral muscles during unilateral movement.

Mapped spinal activity can be used to identify different phases of movement and has been previously used to identify specific phases of gait (*Ivanenko et al., 2006*), (*Santuz et al., 2018*). During hip movement by the control participants, the mapped spinal activity had distinct flexion and extension phases. However, for the SCI participants, we observed only a single phase at the onset of movement, representing either flexion or extension. Moreover, during ankle movement, the mapped activity of the SCI participants was not as broadly distributed as that of the control participants as a result of co-contraction of the distal lower limb muscles.

The lower complexity of muscle activation measured using HFD in the presence of stimulation appeared to be a good marker of proper movement control, as we have observed lower complexity in the muscles directly associated with the BMCA tasks. Although a decrease in the complexity of muscle activation was observed for the SCI participants with stimulation, this decrease occurred independent of task during unilateral movement. This result suggests that the involuntary contraction of contralateral muscles that occurs during stimulation decreases the complexity on the contralateral side and makes it harder to distinguish the differences between the muscle activation complexities of the ipsilateral and contralateral sides. The similarity in HFD complexity between the contralateral and ipsilateral sides during unilateral movement could be used as a measure to quantify a SCI participant’s lack of precise motor control.

During volitional movement in SCI participants with stimulation, a large activation of muscle activity was measured with sEMG, but the movement was not necessarily smooth due to a lack of coordination. Clear isolation of the agonistic and antagonistic muscle activations during extension and flexion can be seen in the bottom right panels of Figure 2, which depict right ankle movement in control participants. In SCI participants, even on their final visit with stimulation (shown in the middle bottom panels), the antagonistic muscles are coactivated together with the agonistic muscles, which results in erratic joint movement. Furthermore, the contralateral muscles are activated involuntarily, showing poor isolation of muscle activation between the left and right sides. Therefore, our future studies will focus on identifying epidural stimulation parameters that improve control to generate more precise limb biomechanics.

### Neural basis of muscle synergies

There is an ongoing debate about whether muscle synergies explain neural changes or task-related changes. In individuals with neurological conditions such as stroke, cerebral palsy, and SCI, the number and structure of muscle synergies changed with the repetition of a single task. This result supports the neural basis of synergies, but this evidence is correlational (*Bizzi and Cheung, 2013*), (*Singh et al., 2018*). Moreover, in individuals without movement disorders, the muscle synergies remained similar when the same tasks were repeated and changed only when the task changed, supporting the task-related hypothesis (*Torres-Oviedo and Ting, 2010*). Studies in intact participants have been unable to resolve the competition between these two hypotheses because the afferent drive through intact ascending tracts is modulated when the task is changed; therefore, there is always a neural component to task-based synergies that cannot be isolated (*Torres-Oviedo and Ting, 2010*), (*Kutch and Valero-Cuevas, 2012*), (*Singh et al., 2020a*), (*Singh et al., 2020b*). However, in the current study, the measurement of changes in muscle synergies under epidural stimulation without the restoration of sensory feedback allows us to isolate the neural effects from the task effects in muscle synergies.

Previous studies have used spinal transection and stimulation in animals to test a direct relationship between changes in neural control and muscle synergies (*Bizzi et al., 1991*), (*Tresch et al., 1999*), (*Saltiel et al., 2001*), (*Saltiel et al., 2005*). Our study is the first to show changes in muscle synergies over time with neuromodulation in SCI participants, as we tested changes in neural drive directly through stimulation. Therefore, our study adds further support for the neural basis of muscle synergy in humans.

Our results strengthen the evidence for the neural basis of muscle synergies in two ways. First, the high numbers of synergies in SCI participants in the absence of stimulation during BMCA tasks are immediately reduced upon stimulation. This reduction in the number of muscle synergies in the absence of a neural controller is counter to the results of (*Kutch and Valero-Cuevas, 2012*) and suggests a non-neural origin of muscle synergies (*Kutch and Valero-Cuevas, 2012*). If synergies were task-related and not neurally controlled, restoring volitional control would not be expected to reduce the number of synergies. The second line of evidence in support of the neural basis of muscle synergies comes from the longitudinal changes observed in this study. A confounding problem in testing the origins of muscle synergies is the adaptation of the CNS to different tasks that reshapes the number and structure of muscle synergies via sensory feedback (*Sawers et al., 2015*), (*Singh et al., 2020a*), (*Singh et al., 2020b*). However, in this study, we were able to measure changes in synergy in the same task performed during the recovery of participants without the restoration of sensory information to the supraspinal region, as most participants were not able to perform the BMCA task without stimulation. This finding suggests that motor learning-based changes in synergies were precluded; however, it is possible, though unlikely, that the changes result from visual-motor learning. Over the follow-up visits, we observed a decrease in the number of synergies, indicating an improvement in muscle synergy due to neural control that was independent of task control. This finding is consistent with those of previous studies showing changes in muscle synergies due to neural changes in development (*Lacquaniti et al., 2012*), (*Dominici et al., 2011*), (*Bizzi and Cheung, 2013*).

### Functional relevance of muscle synergies

Several studies have concluded that three to five muscle synergies are sufficient to account for the basic patterns of muscle activation in the upper and lower limbs (*Ivanenko et al., 2004*), (*Singh et al., 2019*), (*Abd et al., 2021a*). In our study, the control participants had four muscle synergies on average for each of the BMCA tasks. We observed that the SCI participants required all 10 EMG channels (no synergy) at the beginning of the study, and the number of synergies progressively decreased to 4 or 5 by the final session. While the SCI participants achieved similar numbers of muscle synergies as the control participants, there were significant structural differences in their synergies, which we generally observed during the motor control task. An asymmetry in muscle synergies during bilateral movements indicates improper inter-leg coordination and has been previously observed in SCI participants (*Cheung et al., 2012*), (*Zehr et al., 2016*). It is clear from our results that persistent tonic stimulation over many months restored muscle synergies across SCI participants, suggesting plasticity in the spinal circuitry.

### Potential neural mechanism of muscle synergies

Here we put forward a hypothesis for how eSCS restores neural synergies, as illustrated in Figure 10. In control participants, the ascending and descending tracts are intact. During volitional movements, the intact ascending drives act as a task-specific input to the cerebellum, where a predefined model bearing primitive and learned motor behavior exists (*Dominici et al., 2011*). The ascending tracts bring proprioceptive information to the cerebellum, where it is compared with existing predefined models for a specific movement. If an error is detected, the movement is corrected by a complex neural network that includes the cerebellum, cerebral cortex, and basal ganglia. This network assists in the modulation of the descending drives. These modulated descending drives then recruit the necessary muscle synergies to correct the movement.

**Figure 10:**
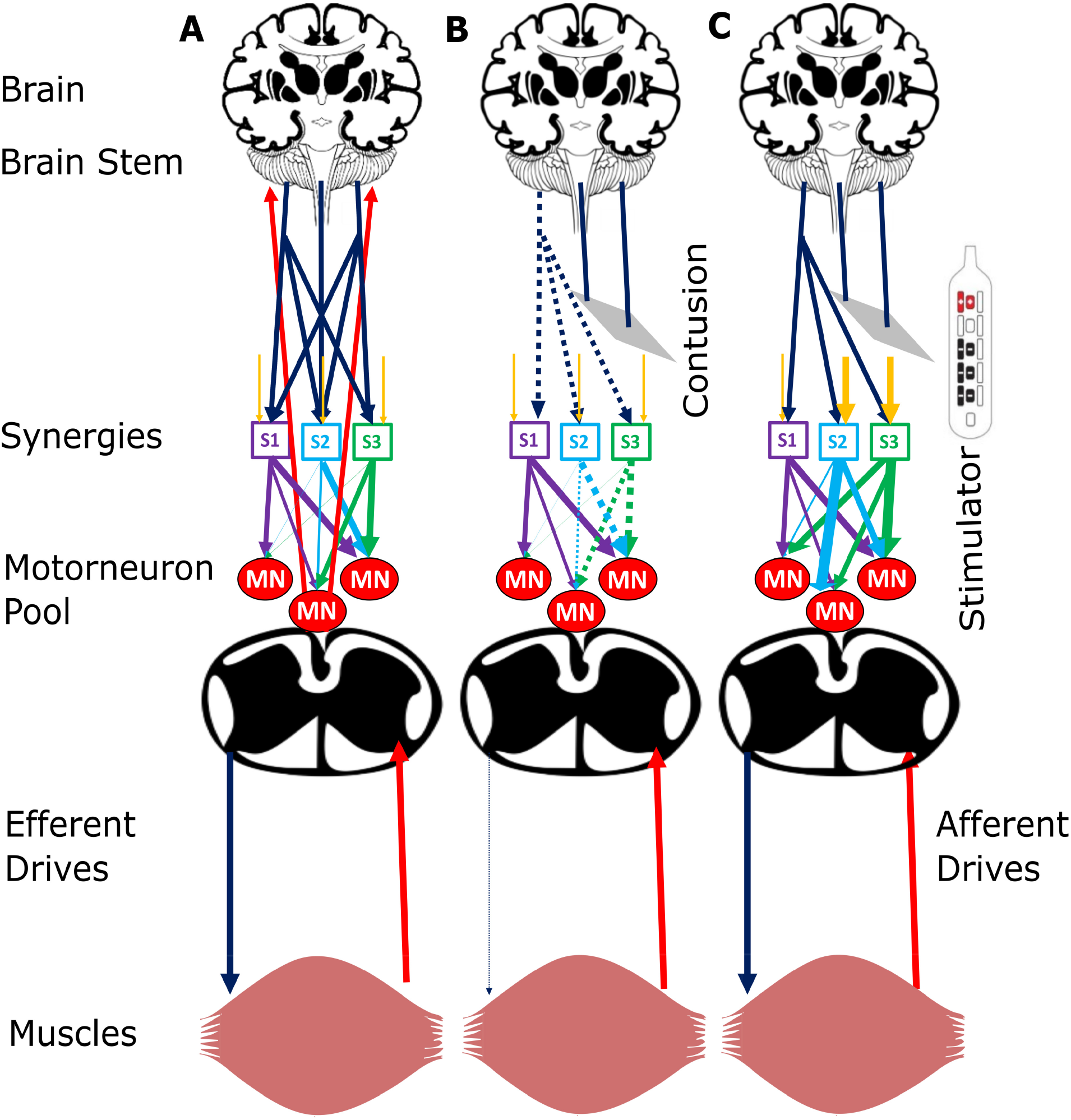
Neural Mechanism. A) Neural circuits of intact participants with efferent drives projecting as a network on muscle synergies (S1, S2, S3). The synergies encode the information of the motor neuronal pool (MN) and activate specific groups of muscles to cause movement. The afferent drives, shown as red arrows, bring information from the task to the supraspinal centers, such as the cerebellum and motor cortex, that reduce any error in movement by modulating the efferent drives. B) Disruption in the spinal circuits after injury obstructs the descending/ascending pathways, causing the inactivation of synergies, which leads to paraplegia. C) Epidural stimulation modulates the sensory afferent and motor efferent drives within the local spinal circuitry, thereby activating muscle synergies and restoring voluntary movement. Sensory feedback to the supraspinal region is absent in SCI participants. The yellow descending arrows represent local spinal circuitry neural drives, the blue descending arrows represent efferent drives, the multicolored descending arrows between synergies and motor neuron pool represent neural network, and ascending red arrows from a muscle represent afferent drives

In our study, the SCI participants were motor and sensory complete, and thus, no sensory information ascended to the supraspinal region. The motor commands from the corticospinal tracts may be severed, but even if a small portion of the spinal cord is left intact, limited cortical signals may project to the spinal cord. The significant reduction in cortical input and drive makes the local spinal circuit unresponsive to these remaining signals, as illustrated in Figure 10b. eSCS modulates the neural drives within the local spinal circuit so that they become sensitive to these limited cortical or supraspinal signals, which then restores some volitional control to the muscles. In addition, it is also possible that the stimulator modulates the residual corticospinal pathways in the dorsal column rather than actual anterolateral and ventral pathways (*Rowald et al., 2022*). These modulated signals further recruit specific muscle synergies to move the limb during a volitional task, as shown in Figure 10c, thus restoring the muscle synergies and voluntary motor control over time.

While Figures 8 and 9 show that muscle synergies are restored in SCS participants, synergies between the sides are not. When the control participants perform bilateral hip and ankle tasks, their first muscle synergy includes muscles from both the right and left sides, indicating synergy among muscles within a leg as well as synergies between legs. In contrast, the right and left leg muscle synergies are isolated into two groups during hip movement and ankle movement by the SCI participants, indicating a lack of inter-leg coordination. We hypothesize that muscle synergies within a leg occur within the spinal cord, which are restored with SCS, but synergies between the legs may be more dependent on supraspinal areas, which are not restored, resulting in asymmetric muscle synergy recruitment during bilateral movement.

## Limitations

One of the limitations of our study is that we did not record the hip extensor muscle groups, which prohibits the analysis of muscle synergies associated with hip extension. The supine posture of the participants made it difficult to record sEMG signals from the hamstrings and gluteus maximus. While sEMG recordings from these muscles would have been useful in the synergy analysis, they were not used because of the high likelihood of the electrodes coming off during the experiment and the noisiness of the signals during BMCA tasks due to movement artifacts.

EMG data were recorded over several sessions to estimate changes in the synergies over time, but because new electrodes were utilized at every follow-up visit, normalization of the amplitudes was needed to compare the muscle activity across sessions. A common approach is to normalize to the maximum voluntary contraction. However, in this study, we were not able to record the maximum voluntary contraction due to the participants’ injuries. Instead, we normalized to the maximum activation of each muscle across all tasks. Previous studies have reported that muscle synergy structures remain consistent across different normalization methods (*Kieliba et al., 2018*). While normalization to the maximum voluntary contraction would be ideal, we believe that the muscle loadings within synergies estimated from the sEMG normalized to the maximum activity on each day is an acceptable alternative and is unlikely to affect the findings of this study.

## Conclusion

In this study, we used muscle complexity and synergy analyses to understand the acute and long-term effects of SCS in participants with chronic motor/sensory complete SCI. Stimulation decreased the muscle activation and localized muscle activity to the rostro-caudal spinal column. It also decreased the muscle complexity, which was measured using an HDF complexity analysis. Thus, SCS changes the AIS score from A to C due to the appearance of motor function below the level of injury, as observed in the synergy and complexity analyses.

We also observed changes in the coordination of muscle groups over time. The number of muscle synergies decreased over the course of the follow-up sessions, and at the end of 13 visits, the number of synergies required to describe 85% of the muscle activation matched that of the control participants. While the number of synergies between the SCI participants with stimulation and the control participants were similar, the muscle loadings within the synergies of the SCI participants did not match those of the control participants. Particularly, when tasks required bilateral movement, the control participants had bilateral muscle synergies whereas the SCI participants did not. Overall, our results suggest that epidural stimulation improves movement control by changing the structure and dimensionality of the muscle synergies via acute and chronic neuromodulation.

Finally, this study provided an opportunity to test in humans whether muscle synergies have a neural or task-dependent basis. The restoration of synergies in the same participant performing the same task over time supports the hypothesis that muscle synergies have a neural basis rather than a task basis.

## Supporting information

Spinal Map Chart

## Data Availability

All data produced in the present study are available upon reasonable request to the corresponding authors

## Author contributions

The study was conceived and designed by RES, DPD, and TIN. AVK, AMP, and US provided input and feedback. AA collected the data. RES, DPD, and TIN performed data analysis and interpretation. RES, DPD, and TIN drafted the article. All authors provided critical revision of the manuscript and final approval of the version to be published.

## Ethical approval

Approval for this study was provided by the institutional review board of both Hennepin County Medical Center. We certify that all applicable institutional and governmental regulations concerning the ethical use of human volunteers were followed during the course of this research.

## References

Abd et al., 2021a. Abd, A. T., Singh, R. E., Iqbal, K., and White, G. (2021a). Investigation of power specific motor primitives in an upper limb rotational motion. J. Mot. Behav., pages 1–12.

Abd et al., 2021b. Abd, A. T., Singh, R. E., Iqbal, K., and White, G. (2021b). A perspective on muscle synergies and different theories related to their adaptation. Biomechanics, 1(2):253–263.

Beck et al., 2020. Beck, L., Veith, D., Linde, M., Gill, M., Calvert, J., Grahn, P., Garlanger, K., Husmann, D., Lavrov, I., Sayenko, D., Strommen, J., Lee, K., and Zhao, K. (2020). Impact of long-term epidural electrical stimulation enabled task-specific training on secondary conditions of chronic paraplegia in two humans. J. Spinal Cord Med., pages 1–6.

Bizzi and Cheung, 2013. Bizzi, E. and Cheung, V. C. K. (2013). The neural origin of muscle synergies. Front. Comput. Neurosci., 7:51.

Bizzi et al., 1991. Bizzi, E., Mussa-Ivaldi, F. A., and Giszter, S. (1991). Computations underlying the execution of movement: a biological perspective. Science, 253(5017):287–291.

Cells, 2017. Cells, G. S. (2017). Stem cell transplantation for spinal cord injuries. https://globalstemcells.com/stem-cell-transplantation-spinal-cord-injuries/.

Cheung et al., 2020. Cheung, V. C. K., Cheung, B. M. F., Zhang, J. H., Chan, Z. Y. S., Ha, S. C. W., Chen, C.-Y., and Cheung, R. T. H. (2020). Plasticity of muscle synergies through fractionation and merging during development and training of human runners. Nat. Commun., 11(1):4356.

Cheung et al., 2005. Cheung, V. C. K., d’Avella, A., Tresch, M. C., and Bizzi, E. (2005). Central and sensory contributions to the activation and organization of muscle synergies during natural motor behaviors. J. Neurosci., 25(27):6419–6434.

Cheung and Seki, 2021. Cheung, V. C. K. and Seki, K. (2021). Approaches to revealing the neural basis of muscle synergies: a review and a critique. J. Neurophysiol., 125(5):1580–1597.

Cheung et al., 2012. Cheung, V. C. K., Turolla, A., Agostini, M., Silvoni, S., Bennis, C., Kasi, P., Paganoni, S., Bonato, P., and Bizzi, E. (2012). Muscle synergy patterns as physiological markers of motor cortical damage. Proc. Natl. Acad. Sci. U. S. A., 109(36):14652–14656.

Courtine et al., 2009. Courtine, G., Gerasimenko, Y., van den Brand, R., Yew, A., Musienko, P., Zhong, H., Song, B., Ao, Y., Ichiyama, R. M., Lavrov, I., Roy, R. R., Sofroniew, M. V., and Edgerton, V. R. (2009). Transformation of nonfunctional spinal circuits into functional states after the loss of brain input. Nat. Neurosci., 12(10):1333–1342.

Craven et al., 2017. Craven, B. C., Giangregorio, L. M., Alavinia, S. M., Blencowe, L. A., Desai, N., Hitzig, S. L., Masani, K., and Popovic, M. R. (2017). Evaluating the efficacy of functional electrical stimulation therapy assisted walking after chronic motor incomplete spinal cord injury: effects on bone biomarkers and bone strength. J. Spinal Cord Med., 40(6):748–758.

Darrow et al., 2019. Darrow, D., Balser, D., Netoff, T. I., Krassioukov, A., Phillips, A., Parr, A., and Samadani, U. (2019). Epidural spinal cord stimulation facilitates immediate restoration of dormant motor and autonomic supraspinal pathways after chronic neurologically complete spinal cord injury. J. Neurotrauma, 36(15):2325–2336.

d’Avella and Bizzi, 2005. d’Avella, A. and Bizzi, E. (2005). Shared and specific muscle synergies in natural motor behaviors. Proceedings of the National Academy of Sciences, 102(8):3076–3081.

Dominici et al., 2011. Dominici, N., Ivanenko, Y. P., Cappellini, G., d’Avella, A., Mondı, V., Cicchese, M., Fabiano, A., Silei, T., Di Paolo, A., Giannini, C., Poppele, R. E., and Lacquaniti, F. (2011). Locomotor primitives in newborn babies and their development. Science, 334(6058):997–999.

Eisdorfer et al., 2020. Eisdorfer, J. T., Smit, R. D., Keefe, K. M., Lemay, M. A., Smith, G. M., and Spence, A. J. (2020). Epidural electrical stimulation: A review of plasticity mechanisms that are hypothesized to underlie enhanced recovery from spinal cord injury with stimulation. Front. Mol. Neurosci., 13:163.

Formento et al., 2018. Formento, E., Minassian, K., Wagner, F., Mignardot, J. B., Le Goff-Mignardot, C. G., Rowald, A., Bloch, J., Micera, S., Capogrosso, M., and Courtine, G. (2018). Electrical spinal cord stimulation must preserve proprioception to enable locomotion in humans with spinal cord injury. Nat. Neurosci., 21(12):1728–1741.

Harkema et al., 2011. Harkema, S., Gerasimenko, Y., Hodes, J., Burdick, J., Angeli, C., Chen, Y., Ferreira, C., Willhite, A., Rejc, E., Grossman, R. G., and Edgerton, V. R. (2011). Effect of epidural stimulation of the lumbosacral spinal cord on voluntary movement, standing, and assisted stepping after motor complete paraplegia: a case study. Lancet, 377(9781):1938–1947.

Ivanenko et al., 2004. Ivanenko, Y. P., Poppele, R. E., and Lacquaniti, F. (2004). Five basic muscle activation patterns account for muscle activity during human locomotion. J. Physiol., 556(Pt 1):267–282.

Ivanenko et al., 2006. Ivanenko, Y. P., Poppele, R. E., and Lacquaniti, F. (2006). Spinal cord maps of spatiotemporal alpha-motoneuron activation in humans walking at different speeds. J. Neurophysiol., 95(2):602–618.

Jones et al., 2014. Jones, M. L., Evans, N., Tefertiller, C., Backus, D., Sweatman, M., Tansey, K., and Morrison, S. (2014). Activity-based therapy for recovery of walking in individuals with chronic spinal cord injury: results from a randomized clinical trial. Arch. Phys. Med. Rehabil., 95(12):2239–46.e2.

Kendall et al., 1993. Kendall, F. P., McCreary, E. K., Provance, P. G., and Others (1993). Muscles: Testing and function. 239 baltimore. MD: Lippincott Williams & Wilkins, 240.

Kieliba et al., 2018. Kieliba, P., Tropea, P., Pirondini, E., Coscia, M., Micera, S., and Artoni, F. (2018). How are muscle synergies affected by electromyography Pre-Processing? IEEE Trans. Neural Syst. Rehabil. Eng., 26(4):882–893.

Kirshblum et al., 2004. Kirshblum, S., Millis, S., McKinley, W., and Tulsky, D. (2004). Late neurologic recovery after traumatic spinal cord injury. Arch. Phys. Med. Rehabil., 85(11):1811–1817.

Kirshblum et al., 2011. Kirshblum, S. C., Burns, S. P., Biering-Sorensen, F., Donovan, W., Graves, D. E., Jha, A., Johansen, M., Jones, L., Krassioukov, A., Mulcahey, M. J., Schmidt-Read, M., and Waring, W. (2011). International standards for neurological classification of spinal cord injury (revised 2011). J. Spinal Cord Med., 34(6):535–546.

Kumar et al., 2018. Kumar, R., Lim, J., Mekary, R. A., Rattani, A., Dewan, M. C., Sharif, S. Y., Osorio-Fonseca, E., and Park, K. B. (2018). Traumatic spinal injury: Global epidemiology and worldwide volume. World Neurosurgery, 113:e345–e363.

Kutch and Valero-Cuevas, 2012. Kutch, J. J. and Valero-Cuevas, F. J. (2012). Challenges and new approaches to proving the existence of muscle synergies of neural origin. PLoS Comput. Biol., 8(5):e1002434.

Lacquaniti et al., 2012. Lacquaniti, F., Ivanenko, Y. P., and Zago, M. (2012). Development of human locomotion. Curr. Opin. Neurobiol., 22(5):822–828.

Lam et al., 2007. Lam, T., Eng, J. J., Wolfe, D. L., Hsieh, J. T., Whittaker, M., and the SCIRE Research Team (2007). A systematic review of the efficacy of gait rehabilitation strategies for spinal cord injury. Top. Spinal Cord Inj. Rehabil., 13(1):32–57.

Lee and Seung, 1999. Lee, D. D. and Seung, H. S. (1999). Learning the parts of objects by non-negative matrix factorization. Nature, 401(6755):788–791.

Marquez-Chin and Popovic, 2020. Marquez-Chin, C. and Popovic, M. R. (2020). Functional electrical stimulation therapy for restoration of motor function after spinal cord injury and stroke: a review. Biomed. Eng. Online, 19(1):34.

Müller et al., 2017. Müller, W., Jung, A., and Ahammer, H. (2017). Advantages and problems of nonlinear methods applied to analyze physiological time signals: human balance control as an example. Scientific Reports, 7(1).

Mushahwar et al., 2007. Mushahwar, V. K., Jacobs, P. L., Normann, R. A., Triolo, R. J., and Kleitman, N. (2007). New functional electrical stimulation approaches to standing and walking. J. Neural Eng., 4(3):S181–97.

Pino et al., 2020. Pino, I. P., Hoover, C., Venkatesh, S., Ahmadi, A., Sturtevant, D., Patrick, N., Freeman, D., Parr, A., Samadani, U., Balser, D., Krassioukov, A., Phillips, A., Netoff, T. I., and Darrow, D. (2020). Long-Term spinal cord stimulation after chronic complete spinal cord injury enables volitional movement in the absence of stimulation. Frontiers in Systems Neuroscience, 14.

Pino et al., 2022. Pino, I. P., Nightingale, T., Hoover, C., Zhao, Z., Cahalan, M., Dorey, T., Walter, M., Soriano, J., Netoff, T., Parr, A., Samadani, U., Phillips, A., Krassioukov, A., and Darrow, D. (2022). The safety of epidural spinal cord stimulation to restore function after spinal cord injury: post-surgical complications and incidence of cardiovascular events. Spinal Cord. Accepted.

Rimini et al., 2017. Rimini, D., Agostini, V., and Knaflitz, M. (2017). Intra-Subject consistency during locomotion: Similarity in shared and Subject-Specific muscle synergies. Front. Hum. Neurosci., 11:586.

Rowald et al., 2022. Rowald, A., Komi, S., Demesmaeker, R., Baaklini, E., Hernandez-Charpak, S. D., Paoles, E., Montanaro, H., Cassara, A., Becce, F., Lloyd, B., Newton, T., Ravier, J., Kinany, N., D’Ercole, M., Paley, A., Hankov, N., Varescon, C., McCracken, L., Vat, M., Caban, M., Watrin, A., Jacquet, C., Bole-Feysot, L., Harte, C., Lorach, H., Galvez, A., Tschopp, M., Herrmann, N., Wacker, M., Geernaert, L., Fodor, I., Radevich, V., Van Den Keybus, K., Eberle, G., Pralong, E., Roulet, M., Ledoux, J.-B., Fornari, E., Mandija, S., Mattera, L., Martuzzi, R., Nazarian, B., Benkler, S., Callegari, S., Greiner, N., Fuhrer, B., Froeling, M., Buse, N., Denison, T., Buschman, R., Wende, C., Ganty, D., Bakker, J., Delattre, V., Lambert, H., Minassian, K., van den Berg, C. A. T., Kavounoudias, A., Micera, S., Van De Ville, D., Barraud, Q., Kurt, E., Kuster, N., Neufeld, E., Capogrosso, M., Asboth, L., Wagner, F. B., Bloch, J., and Courtine, G. (2022). Activity-dependent spinal cord neuromodulation rapidly restores trunk and leg motor functions after complete paralysis. Nat. Med., 28(2):260–271.

Saltiel et al., 2005. Saltiel, P., Wyler-Duda, K., d’Avella, A., Ajemian, R. J., and Bizzi, E. (2005). Localization and connectivity in spinal interneuronal networks: The Adduction–Caudal Extension–Flexion rhythm in the frog. J. Neurophysiol., 94(3):2120–2138.

Saltiel et al., 2001. Saltiel, P., Wyler-Duda, K., D’Avella, A., Tresch, M. C., and Bizzi, E. (2001). Muscle synergies encoded within the spinal cord: evidence from focal intraspinal NMDA iontophoresis in the frog. J. Neurophysiol., 85(2):605–619.

Santuz et al., 2020. Santuz, A., Brüll, L., Ekizos, A., Schroll, A., Eckardt, N., Kibele, A., Schwenk, M., and Arampatzis, A. (2020). Neuromotor dynamics of human locomotion in challenging settings. iScience, 23(1):100796.

Santuz et al., 2018. Santuz, A., Ekizos, A., Eckardt, N., Kibele, A., and Arampatzis, A. (2018). Challenging human locomotion: stability and modular organisation in unsteady conditions. Scientific Reports, 8(1).

Sawers et al., 2015. Sawers, A., Allen, J. L., and Ting, L. H. (2015). Long-term training modifies the modular structure and organization of walking balance control. J. Neurophysiol., 114(6):3359–3373.

Sharrard, 1964. Sharrard, W. J. (1964). THE SEGMENTAL INNERVATION OF THE LOWER LIMB MUSCLES IN MAN. Ann. R. Coll. Surg. Engl., 35:106–122.

Sherwood et al., 1996. Sherwood, A. M., McKay, W. B., and Dimitrijević, M. R. (1996). Motor control after spinal cord injury: assessment using surface EMG. Muscle Nerve, 19(8):966–979.

Singh et al., 2019. Singh, R. E., Iqbal, K., Ullah, S., Alazzawi, A., and others (2019). Gait phase discrimination during kinematically constrained walking on slackline*. 2019 IEEE 15th.

Singh et al., 2020a. Singh, R. E., Iqbal, K., and White, G. (2020a). Proficiency-based recruitment of muscle synergies in a highly perturbed walking task (slackline). Engineering Reports, 2(10):e12253.

Singh et al., 2018. Singh, R. E., Iqbal, K., White, G., and Hutchinson, T. E. (2018). A systematic review on muscle synergies: From building blocks of motor behavior to a neurorehabilitation tool. Appl. Bionics Biomech., 2018:3615368.

Singh et al., 2020b. Singh, R. E., White, G., Delis, I., and Iqbal, K. (2020b). Alteration of muscle synergy structure while walking under increased postural constraints. Cognitive Computation and Systems, 2(2):50–56.

Torres-Oviedo and Ting, 2010. Torres-Oviedo, G. and Ting, L. H. (2010). Subject-specific muscle synergies in human balance control are consistent across different biomechanical contexts. J. Neurophysiol., 103(6):3084–3098.

Tresch et al., 1999. Tresch, M. C., Saltiel, P., and Bizzi, E. (1999). The construction of movement by the spinal cord. Nat. Neurosci., 2(2):162–167.

Zehr et al., 2016. Zehr, E. P., Barss, T. S., Dragert, K., Frigon, A., Vasudevan, E. V., Haridas, C., Hundza, S., Kaupp, C., Klarner, T., Klimstra, M., Komiyama, T., Loadman, P. M., Mezzarane, R. A., Nakajima, T., Pearcey, G. E. P., and Sun, Y. (2016). Neuromechanical interactions between the limbs during human locomotion: an evolutionary perspective with translation to rehabilitation. Exp. Brain Res., 234(11):3059–3081.

Zhao et al., 2021. Zhao, Z., Ahmadi, A., Hoover, C., Grado, L., Peterson, N., Wang, X., Freeman, D., Murray, T., Lamperski, A., Darrow, D., and Netoff, T. I. (2021). Optimization of spinal cord stimulation using bayesian preference learning and its validation. IEEE Trans. Neural Syst. Rehabil. Eng., 29:1987–1997.

