## Supplementary figures and images for "Epidural stimulation restores muscle synergies by modulating neural drives in participants with motor/sensory complete spinal cord injuries"

### Spinal Map.png

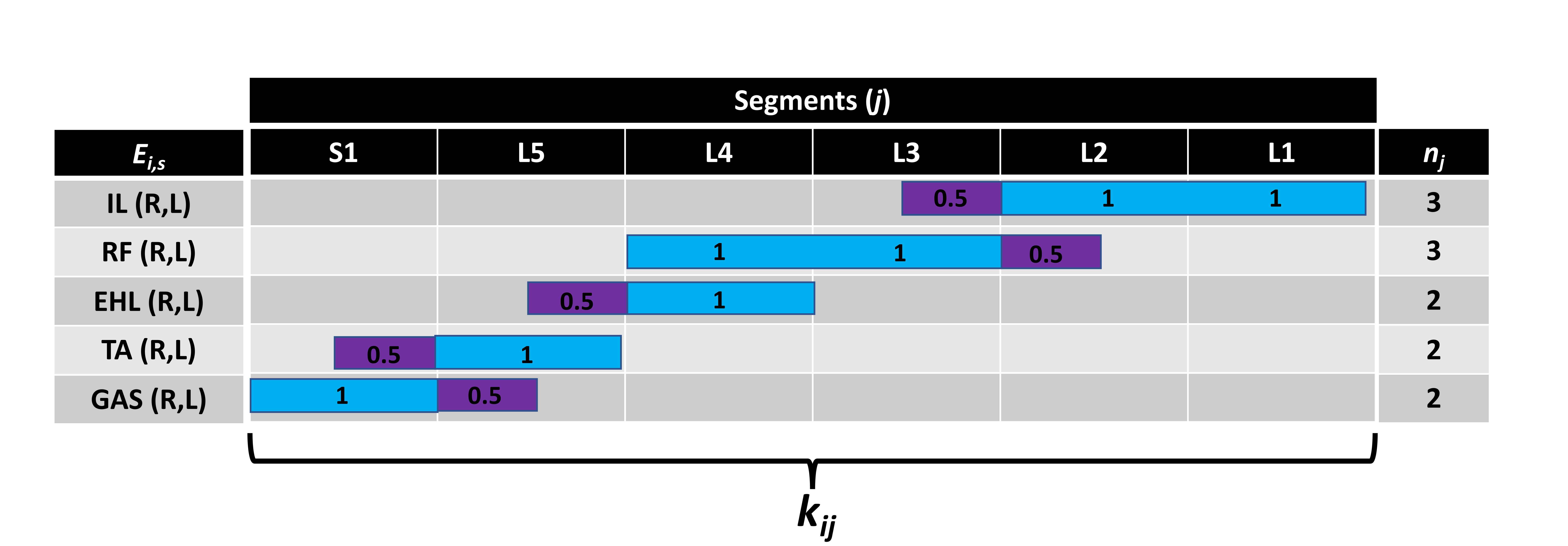
